# The relationship between brain structure and reduced hand dexterity in older adults

**DOI:** 10.1101/2024.11.11.24317033

**Authors:** Anna Manelis, Hang Hu, Skye Satz

**Affiliations:** Department of Psychiatry, University of Pittsburgh, Pittsburgh, PA, USA

**Keywords:** older adults, hand dexterity, Nine Hole Peg Test, MRI, choroid plexus, cortical myelin, neuroinflammation

## Abstract

Hand dexterity is affected by normal aging and neuroinflammatory processes in the brain. Understanding the relationship between hand dexterity and brain structure in neurotypical older adults may inform about prodromal pathological processes, thus providing an opportunity for earlier diagnosis and intervention to improve functional outcomes. This study investigates the associations between hand dexterity and brain measures in neurotypical older adults (≥65 years) using the Nine Hole Peg Test (9HPT) and magnetic resonance imaging (MRI). Elastic net regularized regression revealed that reduced hand dexterity in dominant and non-dominant hands was associated with enlarged volume of the left choroid plexus, the region implicated in neuroinflammatory and altered myelination processes, and reduced myelin content in the left frontal operculum, the region implicated in motor imagery, action production, and higher-order motor functions. Distinct neural mechanisms underlying hand dexterity in dominant and non-dominant hands included the differences in caudate and thalamic volumes as well as altered cortical myelin patterns in frontal, temporal, parietal, and occipital regions supporting sensorimotor and visual processing and integration, attentional control, and eye movements. Although elastic net identified more predictive features for the dominant vs. non-dominant hand, the feature stability was higher for the latter, thus indicating higher generalizability for the non-dominant hand model. Our findings suggest that the 9HPT for hand dexterity may serve as a cost-effective screening tool for early detection of neuroinflammatory and neurodegenerative processes. Longitudinal studies are needed to validate our findings in a larger sample and explore the potential of hand dexterity as an early clinical marker.

## 1. INTRODUCTION

Hand dexterity is the ability to coordinate fine and gross hand and finger movements to perform precise, complex, flexible, and coordinated movements [1]. It was noted that hand dexterity is affected by normal aging [2–4], and neurological disorders like stroke [5–7], multiple sclerosis [8], Parkinson’s [9,10] and Alzheimer’s diseases [11,12]. Importantly, it was noted that reduced hand dexterity might be an early sign of neurological disorders [13,14] and cognitive decline across multiple domains [8]. Understanding the relationship between the changes in hand dexterity and the changes in brain structure may offer insights into prodromal degenerative or neuroinflammatory processes, thus providing an opportunity for earlier diagnosis and intervention to improve functional outcomes.

Action execution is supported by the network of frontal, parietal, temporo-occipital, and subcortical regions [15,16]. Hand movements are controlled by the primary motor cortex [17,18]. The sensory-motor integration, motor planning, and interaction between hand movement and the environment rely on the posterior parietal cortex including the parietal reach region and the anterior intraparietal area [1,19,20]. Hand dexterity requires the interplay between somatosensory, motor [1,21,22], and cognitive function [4,23–25]. It was shown that dexterity in both dominant and non-dominant hands was correlated with memory, executive function, attention, and motor skills [8,26].

Although reduced hand dexterity has been previously observed in individuals with neurodegenerative and neuroinflammatory disorders, only a small number of structural neuroimaging studies have investigated the relationship between hand dexterity and brain anatomy in neurotypical older adults. One recent longitudinal study showed that a decline in hand dexterity was related to increased volume in white matter hyperintensity – brain white matter lesions linked to small vessel disease [7]. Another study pointed to the relationship between cortical myelin content in the primary motor cortex and the temporal precision of finger movements [27]. Altered myelination processes have been linked to neuroinflammation, which, in turn, was associated with choroid plexus volume enlargement [28–31]. Further examination into the relationship between hand dexterity, the choroid plexus volume, and cortical myelin content in the whole brain is needed.

In this study, we examined the relationship between performance on the Nine Hole Peg Test (9HPT), a gold standard for hand dexterity assessment [32–34], and brain structure in neurotypical older adults (≥65 year of age). The brain measures of interest included volumes of subcortical regions implicated in neuroinflammation (e.g., the choroid plexus and lateral ventricles), sensorimotor functions (e.g., the caudate, the thalamus), and cognitive processes (e.g., the hippocampus), as well as cortical myelin levels in the whole brain [35]. We hypothesized that reduced hand dexterity would be associated with enlarged volumes of lateral ventricles and choroid plexus and reduced subcortical regions. Based on one previous study of cortical myelin and hand dexterity [27], we hypothesize that low hand dexterity would be associated with reduced levels of cortical myelin, at least in the regions responsible for motor response. However, based on our previous studies [36,37], an alternative hypothesis is that older adults with lower hand dexterity will demonstrate an imbalance in cortical myelin distribution with abnormal decreases in some cortical regions and abnormal increases in the others.

## 2. MATERIALS AND METHODS

### 2.1 Participants

Twenty right-handed participants took part in this study. The study was approved by the University of Pittsburgh Institutional Review Board (IRB number STUDY20120072) and written informed consent was obtained from all participants who were recruited from the community and the Pitt + Me and Pepper registries. Different aspects of this study as well as inclusion and exclusion strategies have been previously reported by our group [24,38]. All participants were right-handed, fluent in English, had premorbid IQ >85 per the National Adult Reading Test [39] and the Montreal Cognitive Assessment (MoCA) cut-off score >22 [40]. Participants were excluded if they reported having metal in the body, claustrophobia, a history of head injury, neurological disorders, learning disability, current alcohol/drug abuse, and psychotic disorders.

### 2.2 Study procedures

Participants completed cognitive and neurological assessments including visual acuity, cranial nerves, gait, posture, balance, sensation and the Montreal Cognitive Assessment (MoCA) [41] during an in-person office visit. The 9-hole peg test (9HPT) [34,42], the gold standard measurement of hand dexterity, was complete on the day of the scan prior to the scanning procedures. Participants were paid for participation.

#### 2.2.1 Hand dexterity assessment

The 9HPT is used across a wide range of ages and medical conditions [32,43,44]. Participants are instructed to use one hand to take a peg from a dish to place it into each of the 9 holes on the plastic peg board as quickly as possible in any order they choose. After all holes are filled, participants remove the pegs one at a time and place them back into the dish. Time to complete the task is measured in seconds using a stopwatch.

#### 2.2.2. Neuroimaging data acquisition

The neuroimaging data were collected at the University of Pittsburgh/UPMC Magnetic Resonance Research Center using a 3T Siemens Prisma scanner with a 64-channel head coil. High-resolution T1w images were collected using the MPRAGE sequence (TR = 2,400 ms, resolution = 0.8 × 0.8 × 0.8 mm, 208 slices, FOV = 256, TE = 2.22 ms, flip angle = 8°). High-resolution T2w images were collected using TR=3200ms, resolution=0.8x0.8x0.8mm, 208 slices, FOV=256, TE=563ms.Field maps were collected in the AP and posterior-to-anterior (PA) directions using the spin echo sequence (TR = 8,000, resolution = 2 × 2 × 2 mm, FOV = 210, TE = 66 ms, flip angle = 90°, 72 slices). All images were converted to BIDS dataset using *heudiconv*[45] and *dcm2niix* [46].

### 2.3 Data analyses

#### 2.3.1 Demographic and hand dexterity data analyses

The means and standard deviations were computed for demographic and hand dexterity data across all 20 participants. The relationship between demographic, clinical, and hand dexterity variables were examined using linear regression in R (https://www.r-project.org/).

#### 2.3.2 Neuroimaging data processing

Quality of T1w and T2w images was visually inspected and examined using *mriqc version 0.15.1* [47]. Based on this examination, all participants were kept in the structural data analyses.

Structural data were analyzed using Freesurfer v7.4.0 [48]. The structures of interest included bilateral subcortical regions (caudate, putamen, accumbens, hippocampus, and amygdala), lateral ventricles, choroid plexus, cerebral spinal fluid, and white matter hypointensities. The volumes of these structures were adjusted to the estimated total intracranial volume (brain structure volume/ estimated total intracranial volume)*100.

Cortical myelin was calculated based on the T1w/T2w ratio using the minimal preprocessing pipelines developed by the Human Connectome Project [35,49,50] with w*orkbench v1.4.2* and *HCPpipelines-4.1.3* installed on a workstation with GNU/Linux Debian 10 operating system. Bias field correction was performed using a pair of spin echo field maps in *PreFreeSurfer*. The images were registered to standard space using MSMSulc [51] in *PostFreeSurfer*. The 360 region Glasser Atlas [35] was used to parcellate the resulting cortical maps.

#### 2.3.3 Elastic net regression

We used elastic net regularized regression [52,53] to select features (brain structures) that were most important for predicting dominant and non-dominant hand dexterity. Elastic net regularized regression with α=0.5 to control the balance between the ridge and lasso regularizations was conducted across all 20 participants using leave-one-out cross-validation to identify the optimal λ parameter. The predictors included volumes of bilateral subcortical regions (caudate, putamen, pallidum, hippocampus, and amygdala), lateral ventricles, choroid plexus, cerebral spinal fluid, white matter hypointensities, and cortical myelin levels in 360 parcels from Glasser Atlas [35].

After a full-sample regularized regression identified features (brain structures) predicting hand dexterity, we calculated feature stability using a nested cross-validation approach [36,54]. In each loop of nested cross-validation, one participant of 20 was held out and the rest of 19 participants were used in the elastic net regression, which resulted in a total of 20 sparse models. The number of sparse models that selected a feature (brain structure) previously identified by a full-sample elastic net was computed. If a particular brain structure was selected by each of the 20 models, the feature stability was 100%. If only one of the 20 models selected that structure, the feature stability was 5%. A higher feature stability may indicate better result generalizability.

#### 2.3.4 Exploratory analysis

The brain structures selected by the elastic net models for the dominant and non-dominant hands were combined and used in the correlation analysis to determine the strength of the relationships between hand dexterity, demographics (e.g., age, sex), clinical (IQ, MoCA scores, a recent history of falls), and brain measures.

## 3. RESULTS

### 3.1 Demographics and hand dexterity

Participants were eleven women and nine men between 65 and 81 years of age (mean=72±4 years). Their IQ ranged from 97 to 123 (mean =113.9±5.5), and MoCA scores ranged from 23 to 30 (mean=27.6±1.8). Five participants reported a history of falls within the past 12 months.

Hand dexterity was marginally higher in the dominant (right) hand than in the non-dominant (left) hand (dominant: 22±3.4 sec, non-dominant: 23.3±3.4 sec, t(19)= −2.08, p=0.05). The 9HPT scores in the dominant and non-dominant hands were significantly correlated (r=0.66, p=0.002). Hand dexterity in either hand was unrelated to participants’ age, sex, IQ, MOCA scores, and history of falls (all p-values>0.1).

### 3.2 Neuroimaging

The elastic net regularized regression model predicting dexterity in the dominant hand selected 15 brain structures including the left choroid plexus and thalamus, the left 6mp, FOP1, IP0, LBELT, LO1, TGd, VMV3, and the right IFJa, LIPv, STGa, TPOJ2, TPJO3, and V3B. All brain measures except for the left 6pm, IP0, choroid plexus, thalamus, and the right V3B significantly (p<0.05) correlated with the dominant (right) hand performance on the 9HPT. All these features had moderate stability with selection frequency ranging from 25% (when the feature was selected by 5 out of 20 nested models) to 40% (when the feature was selected by 8 out of 20 nested models).

The model predicting dexterity in the non-dominant hand selected five brain structures. Two of these structures, the left choroid plexus and FOP1, overlapped with the structures selected in the dominant hand model. The other three structures included the left IP2, the right caudate, and the right d23ab. All five brain measures strongly and significantly (p<0.05) correlated with the non-dominant (left) hand performance on the 9HPT. All features had higher stability ranging from 45% to 60%.

### 3.3 Exploratory analyses

Participants’ IQ was negatively associated with the right caudate volume (r= −0.58, p=0.007). The right caudate volume (t(18)=-2.26, p=0.036) was larger in participants with a history of falls within the past 12 months compared to those without such history. The MoCA scores positively associated with IQ (r=0.49, p=0.03) and the myelin level in the right TPOJ3 (r=0.5, p=0.03). The myelin level in the right STGa was greater in women, compared to men (t(18)=2.57, p=0.02).

Table 1 reports the brain structures selected by elastic net regularized regression, statistics for the correlation analyses with hand dexterity scores, and feature selection frequency in the nested cross-validation procedure that included 20 models for n-1 (i.e., 19) participants. Figure 1 illustrates all significant correlations (including those between brain measures) as well as the cortical myelin parcels selected by elastic net. For this figure, we combined all brain structures selected in the dominant and non-dominant hand models. Supplemental Materials (Table S1) report a flattened correlation matrix for all correlation results between hand dexterity, demographics, clinical, and brain measures.

**Figure 1.**
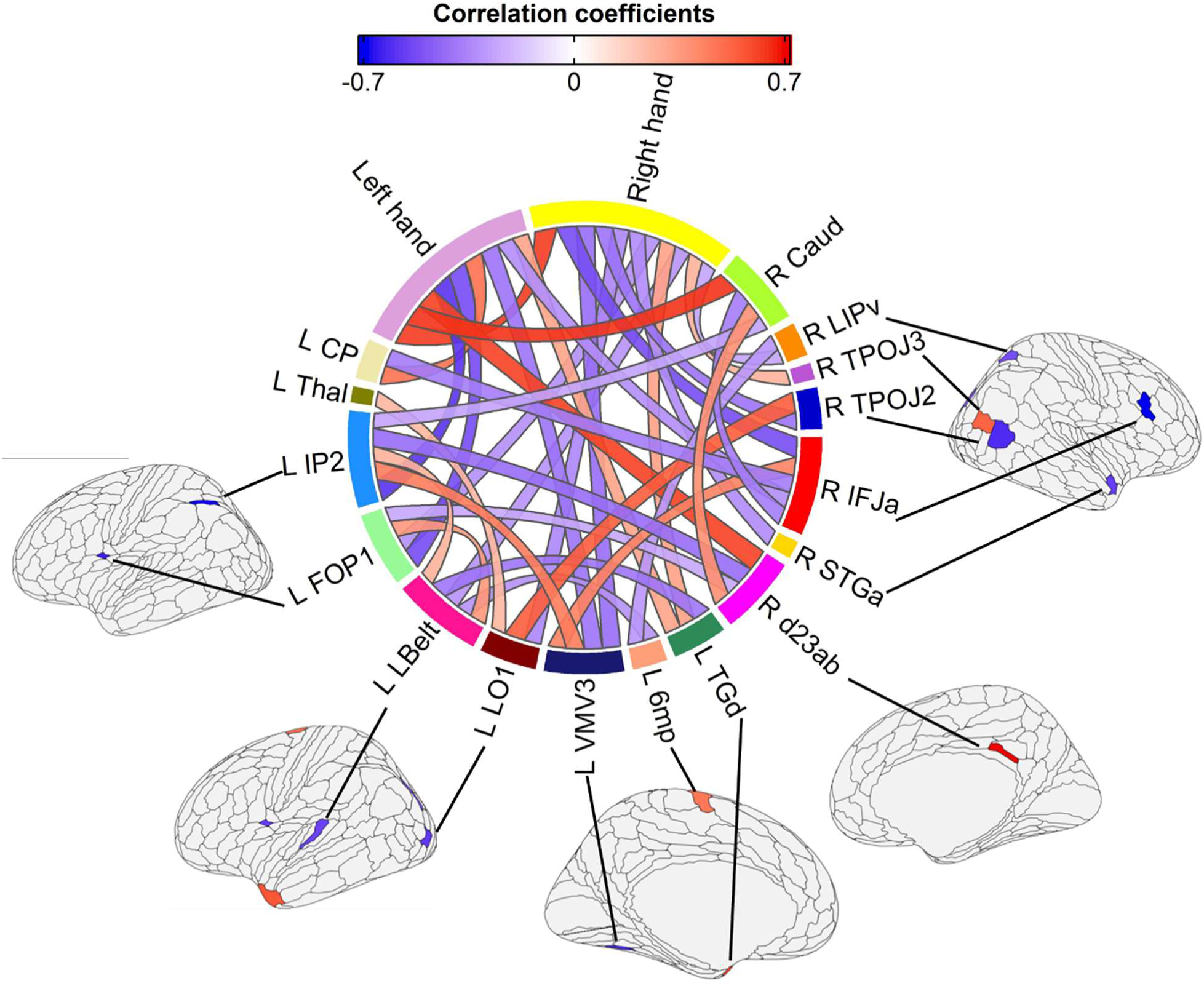
The relationship between a predictor variable (volume and cortical myelin parcels selected by elastic net) and hand dexterity. Brain structures that did not show significant inter-region and dexterity-brain measure relationships are not shown. Cortical myelin parcels selected by elastic net are shown with red (if slower RT on the 9HPT was related to high levels of myelin) and blue (if slower RT on the 9HPT was related to lower levels of myelin) color on the brain

**Table 1.** The correlation between hand dexterity and brain measures in the brain structures selected by elastic net.

| Region | Correlation with hand dexterity | Number of models selected the variable | Feature selection frequency (% of models) |
| --- | --- | --- | --- |
| **Dominant (right) hand dexterity** |  |  |  |
| Left 6mp | $r = 0.4$, $p = 0.08$ | 6 | 30 |
| Left FOP1 | **$r = -0.49$, $p = 0.03$** | 7 | 35 |
| Left IPO | $r = -0.42$, $p = 0.06$ | 5 | 25 |
| Left LBelt | **$r = -0.48$, $p = 0.03$** | 7 | 35 |
| Left LO1 | **$r = -0.51$, $p = 0.02$** | 7 | 35 |
| Left TGD | **$r = 0.49$, $p = 0.03$** | 7 | 35 |
| Left VMV3 | **$r = -0.55$, $p = 0.01$** | 7 | 35 |
| Left choroid plexus | $r = 0.4$, $p = 0.08$ | 5 | 25 |
| Left Thalamus | $r = -0.36$, $p = 0.1$ | 6 | 30 |
| Right IFJa | **$r = -0.6$, $p = 0.005$** | 8 | 40 |
| Right LIPv | **$r = -0.45$, $p = 0.04$** | 7 | 35 |
| Right STGa | **$r = -0.5$, $p = 0.03$** | 7 | 35 |
| Right TPOJ2 | **$r = -0.54$, $p = 0.01$** | 7 | 35 |
| Right TPOJ3 | **$r = 0.46$, $p = 0.04$** | 6 | 30 |
| Right V3B | $r = -0.41, p = 0.07$ | 5 | 25 |
| **Non-dominant (left) hand dexterity** |  |  |  |
| Left FOP1 | $r = -0.61, p = 0.005$ | 11 | 55 |
| Left IP2 | $r = -0.64, p = 0.002$ | 11 | 55 |
| Left.choroid.plexus_etiv | $r = 0.58, p = 0.008$ | 9 | 45 |
| Right d23ab | $r = 0.65, p = 0.002$ | 12 | 60 |
| Right.Caudate_etiv | $r = 0.67, p = 0.001$ | 12 | 60 |

## 4. DISCUSSION

This novel MRI study examined the relationship between dexterity in dominant and non-dominant hands and brain volume and cortical myelin content in neurotypical older adults. Elastic net regularized regression was used to select brain structures important for explaining variability in hand dexterity across all participants. Contrary to previous studies showing reduced dexterity in the non-dominant compared to the dominant hand [33,55], our study revealed only marginally significant differences between dexterity in the dominant and non-dominant hands. We also did not observe dexterity decline with age reported in previous studies [55]. Although dexterity in the dominant and the non-dominant hands correlated, neural underpinnings associated with performance on the 9HPT had overlapping and unique features. So far, only one study examined cortical myelin in the context of hand dexterity and found that myelin content in the motor cortex predicts differences in manual dexterity [27]. Our study extends those findings and shows that hand dexterity is predicted by alterations in the volumes of the choroid plexus, thalamus, and caudate nucleus as well as by cortical myelin content across frontal, temporal, parietal, and occipital cortices.

Reduced dexterity (slower time to complete the 9HPT) in both dominant and non-dominant hands was associated with the left choroid plexus enlargement and the left frontal operculum (FOP1) myelin reduction. Reduced dexterity in the dominant (right) hand was also related to a reduced volume of the left thalamus, reduced cortical myelin content in the frontal (right IFJa), parietal (left IP0, right LIPv), temporal (left LBelt, right STGa), and occipital (left VMV3, right V3B, left LO1) cortices as well as in the right temporal-parietal-occipital junction (TPOJ2), and increased myelin content in the left 6mp (the supplementary motor area), TGd (the temporal polar cortex), and the right TPOJ3 (the temporal-parietal-occipital junction) - the area adjacent to TPOJ2. Reduced dexterity in the non-dominant hand was associated with increased volume of the right caudate, reduced cortical myelin content in the left parietal cortex (IP2), and elevated cortical myelin content in the right posterior cingulate gyrus (d23ab).

The choroid plexus is a brain structure that produces cerebrospinal fluid and controls molecule exchange between the CSF and the bloodstream [56]. Its enlargement was linked to disruption of blood-brain barrier perfusion [28,57], neuroinflammation [28,58,59], altered myelination processes [30], and functional impairments [31,60]. Previous studies observed choroid plexus enlargement in individuals with mood disorders [37], mild cognitive impairment (MCI) [61], Alzheimer’s disease (AD) [60,62], and multiple sclerosis [30,58,63]. The choroid plexus enlargement also predicted conversion from MCI to AD [64]. In our study, the enlarged volume of the choroid plexus was related to reduced myelin content in the contralateral IFJa, the region involved in signal integration across multiple domains, including language and visual non-spatial information [65,66]. This finding highlights the relationship between the choroid plexus volume and altered myelination and points to a potential disruption of the neural signal due to choroid plexus enlargement. Our findings that older adults with reduced hand dexterity also have a larger volume of the choroid plexus suggest that hand dexterity may be an early marker of neuroinflammation in the brain. Considering the cost of MRI examination, administering hand dexterity tests such as the 9HPT once or twice a year might be a low-cost screening tool to identify individuals at risk for neuroinflammation and neurodegeneration disorders. Prospective MRI studies of hand dexterity are needed to understand whether and how the longitudinal trajectory in hand dexterity over several years predicts the trajectory in the choroid plexus volume.

Hand dexterity depends on the participant’s ability to perform precise, complex, flexible, and coordinated hand and finger movements [1]. We found that dexterity in both hands is reduced in those with reduced myelin content in the left FOP1. The left FOP1 is the area in the posterior portions of the inferior frontal gyrus pars opercularis [67]. It is involved in motor imagery, action production, learning motor sequences, and higher-order motor functions [68]. Reduction of myelin content in this region may result in neural signal transmission reduction and decreased ability to coordinate fine motor movements necessary to perform the 9HPT.

Our finding that individuals with lower dexterity in the non-dominant hand had a higher volume of the right caudate was surprising. The caudate nucleus is important for performing purposeful complex fine motor tasks [69]. Previous studies reported this region’s atrophy in MCI, AD, aging [70], and older adults with slower gait speed [71]. Based on these findings, enlargement of the right caudate might be interpreted as brain plasticity to fight neurodegenerative processes. However, our exploratory analyses suggested that a larger volume of caudate nucleus was associated not only with lower non-dominant hand dexterity but also lower IQ and the presence of falls history within the past 12 months. These findings may be indicative of a maladaptive nature of the caudate enlargement as previously reported in autism spectrum disorder [72] and schizophrenia [73] studies.

Elastic net regularized regression selected more brain regions to explain variability in the dominant (15 brain regions), compared to the non-dominant (5 brain regions) hand. However, the feature stability was higher for the non-dominant (45-60%) than the dominant hand (25- 40%), thus suggesting that the results for the non-dominant hand may be more generalizable. Reduced dexterity in the dominant hand was associated with a lower volume of the thalamus and lower cortical myelin content in multiple regions including those implemented in control of attention and eye movements (LIPv) and processing and integrating information from the ventral and dorsal visual streams (VMV3, V3B, LO1). Reduced dexterity in the non-dominant hand was associated with myelin content reduction in the left IP2 which is responsible for sensorimotor integration processes related to fine movements of fingers[74]. Remarkably, the regions in the supplementary motor area, the temporal-parietal-occipital junction, the temporal pole, and the posterior cingulate gyrus had greater cortical myelin content in individuals with lower dexterity. In the context of lower dexterity, these findings might reflect compensatory myelin formation to support fine motor function via a more distributed network of regions or via rerouting information using alternative pathways.

Elastic net regularized regression allowed us to select the most informative features (ROIs) to explain hand dexterity using the most parsimonious model. Considering that a selected set of ROIs comprised less than 5% of all features tested in the model, some features could be removed due to their redundancy. For example, even though the regions like the right IFJa, left TGd, and left VMV3 were selected by the elastic net model for the dominant hand but were not selected by the model for the non-dominant hand, their myelin content still correlated with the non-dominant hand dexterity. In contrast, the regions unique for the non-dominant hand elastic net model, such as the right d23ab and the left IP2, did not correlate with the dominant hand performance. We hypothesize that the non-dominant hand fine movements may be less automatic than the dominant hand movements and engage regions involved in external focus and visuospatial awareness, such as d23ab [75].

One of this study’s limitations is a small sample size, which reduces our ability to generalize these results to the population. Another limitation is a cross-sectional design that did not allow us to examine the relationship between within-subject trajectories in hand dexterity and brain structure. Future longitudinal studies should replicate the result of this study and confirm the utility of using hand dexterity as a proxy for screening of neuroinflammatory and degenerative processes in the brain. While the present study focused on neurotypical older adults, it may also be beneficial to examine participants with MCI to better understand how the relationship between hand dexterity and brain measures might differ for someone with known neurocognitive impairments and whether changes to hand dexterity are indicative of transition from MCI into AD. Recent work revealed that hand dexterity training enhances cognitive abilities in older adults [76], and motor learning promotes increased myelination and supports remyelination within specific critical windows required for effectiveness [77]. Thus, understanding how hand dexterity is related to neuroinflammatory, and degenerative processes might provide intervention targets for earlier detection and intervention, such as physical therapy and dexterity training to help maintain motor and cognitive function.

In conclusion, this study examined the connection between hand dexterity in both dominant and non-dominant hands, brain volume, and cortical myelin content in neurotypical older adults. Hand dexterity was associated with enlarged volume of the choroid plexus, a marker associated with neuroinflammation, and the caudate nucleus and reduced volume of the thalamus and cortical myelin in multiple brain areas supporting sensorimotor processing and integration. Our findings highlight the potential of using simple dexterity tests such as the 9HPT as a cost-effective screening tool for early diagnosis of neuroinflammatory and neurodegenerative processes in the brain, thus offering promising avenues for earlier intervention and prevention strategies.

## Data Availability

All data produced in the present study are available upon reasonable request to the authors

## AUTHOR CONTRIBUTIONS

A.M. – designed the study, curated study development, evaluated data quality, analyzed and interpreted the data, drafted, and critically evaluated the manuscript

H.H. – evaluated data quality, analyzed and interpreted the data, drafted, and critically evaluated the manuscript

S.S. – curated study development, recruited participants, acquired data, evaluated data quality, drafted, and critically evaluated the manuscript

All authors have read and approved the final version of the manuscript and agreed to be accountable for all aspects of this work.

## FUNDING ACKNOWLEDGEMENTS

This work was supported by the Department of Psychiatry at the University of Pittsburgh.

## DISCLOSURE

A.M., H.H., R.M., and S.S. declare no conflict of interest.

## ACKNOWLEDGMENTS

The authors thank participants for taking part in this research study.

## SUPPLEMENTAL MATERIALS

**Table S1.** Correlations between hand dexterity, demographics, clinical, and brain measures.

| Variable 1 | Variable 2 | Correlation coefficient | p-value | Hand for which elastic net model was conducted<br>Dominant hand = right. |
| --- | --- | --- | --- | --- |
|  |  |  |  | **Non-dominant hand = left.** |
| **Correlations between clinical and hand dexterity variables** |  |  |  |  |
| **9HPT dominant hand** | **9HPT non-dominant hand** | **0.66** | **0.002** |  |
| **MoCA** | **IQ** | **0.49** | **0.03** |  |
| Age | IQ | -0.36 | n.s. |  |
| Age | MoCA | -0.27 | n.s. |  |
| 9HPT non-dominant hand | Age | 0.37 | n.s. |  |
| 9HPT non-dominant hand | IQ | -0.19 | n.s. |  |
| 9HPT non-dominant hand | MoCA | 0.02 | n.s. |  |
| 9HPT dominant hand | Age | 0.18 | n.s. |  |
| 9HPT dominant hand | IQ | 0.01 | n.s. |  |
| 9HPT dominant hand | MoCA | 0.24 | n.s. |  |
| **Correlation of clinical and dexterity variables with brain variables** |  |  |  |  |
| **9HPT non-dominant hand** | **Right Caudate** | **0.67** | **0.001** | **left** |
| **9HPT non-dominant hand** | **Left IP2** | **-0.64** | **0.002** | **left** |
| **9HPT non-dominant hand** | **Right d23ab** | **0.65** | **0.002** | **left** |
| **9HPT non-dominant hand** | **Left FOP1** | **-0.61** | **0.005** | **both** |
| **9HPT dominant hand** | **Right IFJa** | **-0.6** | **0.005** | **right** |
| **IQ** | **Right Caudate** | **-0.58** | **0.007** | **left** |
| **Age** | **Left IP2** | **-0.57** | **0.008** | **left** |
| **9HPT non-dominant hand** | **Left choroid plexus** | **0.58** | **0.008** | **both** |
| **Age** | **Left LBelt** | **-0.56** | **0.01** | **right** |
| **9HPT dominant hand** | **Left VMV3** | **-0.55** | **0.01** | **right** |
| **9HPT non-dominant hand** | **Left VMV3** | **-0.54** | **0.01** | **right** |
| **9HPT dominant hand** | **Right TPOJ2** | **-0.54** | **0.01** | **right** |
| **9HPT dominant hand** | **Left LO1** | **-0.51** | **0.02** | **right** |
| **MoCA** | **Right TPOJ3** | **0.5** | **0.03** | **right** |
| **9HPT dominant hand** | **Right STGa** | **-0.5** | **0.03** | **right** |
| **9HPT non-dominant hand** | **Right IFJa** | **-0.49** | **0.03** | **right** |
| **9HPT dominant hand** | **Left FOP1** | **-0.49** | **0.03** | **both** |
| **9HPT dominant hand** | **Left TGd** | **0.49** | **0.03** | **right** |
| **9HPT non-dominant hand** | **Left TGd** | **0.48** | **0.03** | **right** |
| **9HPT dominant hand** | **Left LBelt** | **-0.48** | **0.03** | **right** |
| **9HPT dominant hand** | **Right LIPv** | **-0.45** | **0.04** | **right** |
| **9HPT dominant hand** | **Right TPOJ3** | **0.46** | **0.04** | **right** |
| Age | Left 6mp | 0.09 | n.s. | right |
| Age | Left FOP1 | -0.22 | n.s. | both |
| Age | Left IPo | 0.19 | n.s. | right |
| Age | Left LO1 | -0.13 | n.s. | right |
| Age | Left TGd | 0.11 | n.s. | right |
| Age | Left VMV3 | -0.21 | n.s. | right |
| Age | Left choroid plexus | 0.18 | n.s. | both |
| Age | Left Thalamus | -0.44 | n.s. | right |
| Age | Right d23ab | 0.23 | n.s. | left |
| Age | Right IFJa | -0.17 | n.s. | right |
| Age | Right LIPv | -0.17 | n.s. | right |
| Age | Right STGa | 0.11 | n.s. | right |
| Age | Right TPOJ2 | -0.18 | n.s. | right |
| Age | Right TPOJ3 | -0.44 | n.s. | right |
| Age | Right V3B | -0.2 | n.s. | right |
| Age | Right Caudate | 0.44 | n.s. | left |
| IQ | Left 6mp | 0.13 | n.s. | right |
| IQ | Left FOP1 | 0.05 | n.s. | both |
| IQ | Left IPo | -0.15 | n.s. | right |
| IQ | Left IP2 | 0.4 | n.s. | left |
| IQ | Left LBelt | 0.07 | n.s. | right |
| IQ | Left LO1 | -0.05 | n.s. | right |
| IQ | Left TGD | 0.21 | n.s. | right |
| IQ | Left VMV3 | 0.24 | n.s. | right |
| IQ | Left choroid plexus | 0.36 | n.s. | both |
| IQ | Left Thalamus | -0.27 | n.s. | right |
| IQ | Right d23ab | -0.3 | n.s. | left |
| IQ | Right IFJa | 0.1 | n.s. | right |
| IQ | Right LIPv | -0.12 | n.s. | right |
| IQ | Right STGa | 0.11 | n.s. | right |
| IQ | Right TPOJ2 | 0.01 | n.s. | right |
| IQ | Right TPOJ3 | 0.36 | n.s. | right |
| IQ | Right V3B | 0.18 | n.s. | right |
| 9HPT non-dominant hand | Left 6mp | 0.02 | n.s. | right |
| 9HPT non-dominant hand | Left IPo | -0.05 | n.s. | right |
| 9HPT non-dominant hand | Left LBelt | -0.42 | n.s. | right |
| 9HPT non-dominant hand | Left LO1 | -0.19 | n.s. | right |
| 9HPT non-dominant hand | Left Thalamus | -0.22 | n.s. | right |
| 9HPT non-dominant hand | Right LIPv | 0.13 | n.s. | right |
| 9HPT non-dominant hand | Right STGa | -0.22 | n.s. | right |
| 9HPT non-dominant hand | Right TPOJ2 | -0.3 | n.s. | right |
| 9HPT non-dominant hand | Right TPOJ3 | 0.29 | n.s. | right |
| 9HPT non-dominant hand | Right V3B | -0.24 | n.s. | right |
| MoCA | Left 6mp | 0.08 | n.s. | right |
| MoCA | Left FOP1 | 0.05 | n.s. | both |
| MoCA | Left IPo | -0.22 | n.s. | right |
| MoCA | Left IP2 | 0.39 | n.s. | left |
| MoCA | Left LBelt | 0 | n.s. | right |
| MoCA | Left LO1 | -0.08 | n.s. | right |
| MoCA | Left TGD | 0.04 | n.s. | right |
| MoCA | Left VMV3 | -0.13 | n.s. | right |
| MoCA | Left choroid plexus | 0.43 | n.s. | both |
| MoCA | Left Thalamus | 0.2 | n.s. | right |
| MoCA | Right d23ab | 0.03 | n.s. | left |
| MoCA | Right IFJa | -0.42 | n.s. | right |
| MoCA | Right LIPv | -0.25 | n.s. | right |
| MoCA | Right STGa | 0.02 | n.s. | right |
| MoCA | Right TPOJ2 | -0.23 | n.s. | right |
| MoCA | Right V3B | 0.33 | n.s. | right |
| MoCA | Right.Caudate_etiv | -0.04 | n.s. | left |
| 9HPT dominant hand | Left 6mp | 0.4 | n.s. | right |
| 9HPT dominant hand | Left IPo | -0.42 | n.s. | right |
| 9HPT dominant hand | Left IP2 | -0.39 | n.s. | left |
| 9HPT dominant hand | Left choroid plexus | 0.4 | n.s. | both |
| 9HPT dominant hand | Left Thalamus | -0.36 | n.s. | right |
| 9HPT dominant hand | Right d23ab | 0.23 | n.s. | left |
| 9HPT dominant hand | Right V3B | -0.41 | n.s. | right |
| 9HPT dominant hand | Right.Caudate_etiv | 0.37 | n.s. | left |
| **Correlation between brain variables** |  |  |  |  |
| **Right TPOJ2** | **Left LO1** | **0.61** | **0.004** | **right** |
| **Right IFJa** | **Left VMV3** | **0.55** | **0.01** | **right** |
| **Right d23ab** | **Left IP2** | **-0.55** | **0.01** | **left** |
| **Left TGd** | **Left LBelt** | **-0.54** | **0.02** | **right** |
| **Left choroid plexus** | **Right IFJa** | **-0.52** | **0.02** | **right** |
| **Right Caudate** | **Right d23ab** | **0.52** | **0.02** | **left** |
| **Left FOP1** | **Left LBelt** | **0.51** | **0.02** | **right** |
| **Left 6mp** | **Left LBelt** | **-0.49** | **0.03** | **right** |
| **Right LIPv** | **Left 6mp** | **-0.48** | **0.03** | **right** |
| **Right Caudate** | **Left IP2** | **-0.45** | **0.04** | **left** |
| **Right d23ab** | **Left FOP1** | **-0.45** | **0.05** | **left** |
| **Left Thalamus** | **Left LO1** | **0.45** | **0.05** | **right** |
| Left 6mp | Left FOP1 | -0.27 | n.s. | right |
| Left 6mp | Left IPo | -0.26 | n.s. | right |
| Left 6mp | Left TGd | 0.09 | n.s. | right |
| Left 6mp | Left VMV3 | -0.42 | n.s. | right |
| Left FOP1 | Left IPo | 0.15 | n.s. | right |
| Left FOP1 | Left IP2 | 0.4 | n.s. | left |
| Left FOP1 | Left TGd | -0.44 | n.s. | right |
| Left FOP1 | Left VMV3 | 0.16 | n.s. | right |
| Left IPo | Left LBelt | 0.11 | n.s. | right |
| Left IPo | Left VMV3 | 0.13 | n.s. | right |
| Left LO1 | Left 6mp | -0.28 | n.s. | right |
| Left LO1 | Left FOP1 | 0.05 | n.s. | right |
| Left LO1 | Left IPo | 0.12 | n.s. | right |
| Left LO1 | Left LBelt | 0 | n.s. | right |
| Left LO1 | Left TGd | -0.01 | n.s. | right |
| Left LO1 | Left VMV3 | 0.17 | n.s. | right |
| Left TGd | Left IPo | 0.06 | n.s. | right |
| Left TGd | Left VMV3 | -0.24 | n.s. | right |
| Left VMV3 | Left LBelt | 0.37 | n.s. | right |
| Left choroid plexus | Left 6mp | 0.2 | n.s. | right |
| Left choroid plexus | Left FOP1 | -0.36 | n.s. | both |
| Left choroid plexus | Left IPo | -0.29 | n.s. | right |
| Left choroid plexus | Left IP2 | -0.2 | n.s. | left |
| Left choroid plexus | Left LBelt | -0.33 | n.s. | right |
| Left choroid plexus | Left LO1 | -0.08 | n.s. | right |
| Left choroid plexus | Left TGd | 0.37 | n.s. | right |
| Left choroid plexus | Left VMV3 | -0.25 | n.s. | right |
| Left choroid plexus | Right d23ab | 0.41 | n.s. | left |
| Left choroid plexus | Right LIPv | -0.11 | n.s. | right |
| Left choroid plexus | Right STGa | 0.12 | n.s. | right |
| Left choroid plexus | Right TPOJ2 | -0.01 | n.s. | right |
| Left choroid plexus | Right TPOJ3 | 0.24 | n.s. | right |
| Left choroid plexus | Right V3B | 0 | n.s. | right |
| Left choroid plexus | Right Caudate | 0.37 | n.s. | left |
| Left Thalamus | Left 6mp | -0.31 | n.s. | right |
| Left Thalamus | Left FOP1 | 0.15 | n.s. | right |
| Left Thalamus | Left IPo | 0.22 | n.s. | right |
| Left Thalamus | Left LBelt | 0.42 | n.s. | right |
| Left Thalamus | Left TGD | -0.32 | n.s. | right |
| Left Thalamus | Left VMV3 | 0.08 | n.s. | right |
| Left Thalamus | Left choroid plexus | -0.33 | n.s. | right |
| Left Thalamus | Right IFJa | -0.14 | n.s. | right |
| Left Thalamus | Right LIPv | 0.18 | n.s. | right |
| Left Thalamus | Right STGa | -0.08 | n.s. | right |
| Left Thalamus | Right TPOJ2 | 0.27 | n.s. | right |
| Left Thalamus | Right TPOJ3 | -0.08 | n.s. | right |
| Left Thalamus | Right V3B | 0.3 | n.s. | right |
| Right IFJa | Left 6mp | -0.22 | n.s. | right |
| Right IFJa | Left FOP1 | 0.3 | n.s. | right |
| Right IFJa | Left IPo | 0.4 | n.s. | right |
| Right IFJa | Left LBelt | 0.36 | n.s. | right |
| Right IFJa | Left LO1 | 0.05 | n.s. | right |
| Right IFJa | Left TGD | -0.19 | n.s. | right |
| Right IFJa | Right STGa | 0.13 | n.s. | right |
| Right IFJa | Right TPOJ2 | 0.16 | n.s. | right |
| Right IFJa | Right TPOJ3 | -0.25 | n.s. | right |
| Right LIPv | Left FOP1 | 0.04 | n.s. | right |
| Right LIPv | Left IPo | 0.17 | n.s. | right |
| Right LIPv | Left LBelt | 0.42 | n.s. | right |
| Right LIPv | Left LO1 | 0.24 | n.s. | right |
| Right LIPv | Left TGD | -0.27 | n.s. | right |
| Right LIPv | Left VMV3 | -0.01 | n.s. | right |
| Right LIPv | Right IFJa | 0.28 | n.s. | right |
| Right LIPv | Right STGa | 0.43 | n.s. | right |
| Right LIPv | Right TPOJ2 | 0.08 | n.s. | right |
| Right LIPv | Right TPOJ3 | 0.1 | n.s. | right |
| Right STGa | Left 6mp | 0.01 | n.s. | right |
| Right STGa | Left FOP1 | 0.09 | n.s. | right |
| Right STGa | Left IPo | 0.14 | n.s. | right |
| Right STGa | Left LBelt | 0.13 | n.s. | right |
| Right STGa | Left LO1 | -0.09 | n.s. | right |
| Right STGa | Left TGD | -0.27 | n.s. | right |
| Right STGa | Left VMV3 | 0.09 | n.s. | right |
| Right STGa | Right TPOJ2 | 0.07 | n.s. | right |
| Right STGa | Right TPOJ3 | -0.06 | n.s. | right |
| Right TPOJ2 | Left 6mp | 0.16 | n.s. | right |
| Right TPOJ2 | Left FOP1 | 0.24 | n.s. | right |
| Right TPOJ2 | Left IPo | 0.32 | n.s. | right |
| Right TPOJ2 | Left LBelt | 0.05 | n.s. | right |
| Right TPOJ2 | Left TGD | -0.08 | n.s. | right |
| Right TPOJ2 | Left VMV3 | 0.06 | n.s. | right |
| Right TPOJ2 | Right TPOJ3 | -0.24 | n.s. | right |
| Right TPOJ3 | Left 6mp | 0.28 | n.s. | right |
| Right TPOJ3 | Left FOP1 | -0.13 | n.s. | right |
| Right TPOJ3 | Left IPo | -0.28 | n.s. | right |
| Right TPOJ3 | Left LBelt | 0 | n.s. | right |
| Right TPOJ3 | Left LO1 | -0.32 | n.s. | right |
| Right TPOJ3 | Left TGD | 0.2 | n.s. | right |
| Right TPOJ3 | Left VMV3 | -0.31 | n.s. | right |
| Right V3B | Left 6mp | -0.03 | n.s. | right |
| Right V3B | Left FOP1 | 0.3 | n.s. | right |
| Right V3B | Left IPo | 0.33 | n.s. | right |
| Right V3B | Left LBelt | 0.21 | n.s. | right |
| Right V3B | Left LO1 | 0.04 | n.s. | right |
| Right V3B | Left TGD | -0.12 | n.s. | right |
| Right V3B | Left VMV3 | 0.15 | n.s. | right |
| Right V3B | Right IFJa | 0.21 | n.s. | right |
| Right V3B | Right LIPv | 0.03 | n.s. | right |
| Right V3B | Right STGa | 0.23 | n.s. | right |
| Right V3B | Right TPOJ2 | 0.26 | n.s. | right |
| Right V3B | Right TPOJ3 | 0.13 | n.s. | right |
| Right Caudate | Left FOP1 | -0.37 | n.s. | left |

Note: The abbreviation “n.s.” stands for not significant results ( $p < 0.05$ )

## Notes

### Competing Interest Statement

The authors have declared no competing interest.

### Author Declarations

The study was approved by the University of Pittsburgh Institutional Review Board (IRB number STUDY20120072) and written informed consent was obtained from all participants

